# Evaluating the Association between Concussion Policies, Equity, and Sport-Related Concussion in Children and Youth: a Population-Based Study (2014-2024)

**DOI:** 10.64898/2026.01.15.26344199

**Authors:** Alison Macpherson, Matthew Sudiyono, Carolyn Emery, Stephanie Cowle, Pamela Fuselli, Lavina Matai, Linda Rothman

## Abstract

**Background/aims:** Sport-related concussions (SRCs) are becoming a global public health concern as new research and policies emerge. In the Province of Ontario, a 2014 Policy/Program Memorandum (PPM 158) required all school boards to establish a policy that promoted concussion education, awareness, and tracking. This was followed by Rowan’s Law in 2018 which emphasized sports-related treatments and tracking of concussions. The objective of this study was to examine the association between these concussion policies and emergency department (ED) visits for SRCs in children and youth (5-19) in Ontario by material deprivation from 2014-2024.

**Methods:** This population-based study used routinely collected administrative data from ICES in Ontario, Canada, specifically the National Ambulatory Care Reporting System (NACRS). All ED visits for children and adolescents ages 5-19 with ICD-10 (International Classification of Disease version 10) S060 identifying concussion were included with a corresponding ICD-10 mechanism of injury code related to sports. The number of SRCs and the percent of SRCs within total concussions were analyzed. An interrupted time series analysis was conducted to examine changes in ED visits for SRC from 2014-2018 (the years following the release of PPM 158 up until the release of Rowan’s Law), from 2020-2021 (the years during the COVID-19 pandemic), and in 2019, 2022, 2023, and 2024 (the years following Rowan’s Law).

**Results:** The number of ED visits initially increased after the introduction of Policy/Program Memorandum No. 158 but started to decline after 2018 when Rowan’s Law was introduced. The numbers have remained lower post-COVID-19 pandemic suggesting that this law may have a positive impact. By material deprivation quintile, the highest income quintile saw significantly lower counts of ED (β = −628.7 (p = 0.001)) following concussion laws compared to before.

**Conclusions:** Rowan’s Law appears to have a positive lasting impact on Ontario youths by reducing ED visits for SRCs. Educational resources related to awareness and identification of concussions should continue to be available to children and youth, coaches, and parents.

**What is known on this subject:** Sport-related concussions among youths are associated with serious long-term burdens, with social disparities impacting their access to much-needed care. In Ontario, Canada, a concussion-related policy (PPM 158) and a law (Rowan’s Law) that were adopted in 2014 and 2018, respectively, have yet to be analyzed in terms of its impact.

**What this study adds:** This study contributes to a greater understanding of the association between Rowan’s Law and children and youth seeking care for sports-related concussions in emergency departments. Further, it provides insight into differences in emergency department visits by material deprivation quintiles.

**How might this affect policy, practice and research:** The results of this study may encourage other jurisdictions to enact similar legislation, and can inform school boards and sporting organizations to have an equity-driven lens when delivering concussion education to students, parents, and coaches.

## Introduction

Sports-related concussions represent a pressing policy concern for Canadian youths [1]. Between 2016 and 2017, 46,000 Canadian children aged 5 to 19 had a concussion [2]. This demographic accounts for 70% of all concussions, with 1-in-5 adolescents estimated to sustain a concussion by age 16 [3]. Among these concussion events, half occurred while playing a sport, particularly high-risk sports like rugby and ice hockey [2]. Key mandates including Policy/Program Memorandum No. 158, released in 2014, and Rowan’s Law, enacted in 2018, were introduced to establish guidelines in concussion management [4,5]. These policies focused on prevention through education, identifying concussions, and establishing return-to-school protocols [4,5]. Rowan’s Law specifically targeted sports-related concussions by incorporating coaches, sport staff, parents, and student athletes in concussion education [4,5]. While such Canadian policies have improved the management of concussions, research suggests that existing mandates in Ontario still require better adherence [6,7].

Given that up to 95% of traumatic brain injuries are associated with concussions, poor adherence to guidelines may result in drastic complications such as post-concussion syndrome, post-traumatic stress syndrome, and chronic traumatic encephalopathy [6,8,9]. These ailments carry long-term physical, social, and psychological consequences for children and youth and result in substantial burdens in healthcare and emergency response systems [9]. To illustrate, Ontario emergency departments (ED) report that TBIs have amounted to total lifetime medical and productivity costs of up to ~$1.22 billion, which compound into other unseen societal costs [10]. For youths and young adults between the age ranges of 5-14 and 15-24, these costs increase from $80 million to $130 million [10]. Further, as EDs represent an integral site of concussion surveillance, a growing body of literature has stressed the importance of examining inequities in ED utilization across socioeconomic statuses [1,11]. A systematic review established that most studies reported higher injury risks among those with low socioeconomic status [11]. Another study noted that having public (vs private) insurance influenced return-to-school and return-to-sport timelines among youth [12].

Despite the established importance of linking inequities to concussion outcomes, no study has examined how material deprivation relates to ED visits for sports-related concussions among Canadian youths. Further, no study has assessed how these trends align with the rollout of key Ontario-based mandates or disruptions such as the COVID-19 pandemic. To address this gap, 10 years (2014-2024) of Ontario ED data on SRCs were analyzed using an interrupted time series regression, stratified by material deprivation quartiles. This study will assess the equity implications and the association between of two major concussion policies on ED visits for concussions among children and youth.

## Methods

### Study design and overview

A population-based study was conducted using Ontario ED data from the National Ambulatory Care Reporting System (NACRS) for the fiscal years 2014-2024 [13]. This data is obtained ICES which was originally collected by the Canadian Institute of Health Information [13,14]. In 2018, the institute formerly known as the Institute for Clinical Evaluative Sciences formally adopted the initialism ICES as its official name. This change acknowledges the growth and evolution of the organization’s research since its inception in 1992, while retaining the familiarity of the former acronym within the scientific community and beyond. ICES is an independent, non-profit research institute funded by an annual grant from the Ontario Ministry of Health (MOH) and the Ministry of Long-Term Care (MLTC). As a prescribed entity under Ontario’s privacy legislation, ICESc is authorized to collect and use health care data for the purposes of health system analysis, evaluation and decision support. Secure access to these data is governed by policies and procedures that are approved by the Information and Privacy Commissioner of Ontario. These datasets were linked using unique encoded identifiers and analysed at ICES. Ontario is Canada’s most populous province, and like other Canadian provinces, has universal health care for hospitals, EDs, and most visits to physician offices. The use of the data in this project is authorized under section 45 of Ontario’s Personal Health Information Protection Act (PHIPA) and does not require review by a Research Ethics Board. Patients and the public were not consulted in the conduct of this research. However, Parachute, Canada’s national injury prevention charity and a founding member of the Canadian Concussion Network were involved.

The study population included Ontario children and youth aged 5-19 years old. NARCS captured ED visits of all children and youth who presented to the ED with a diagnosis of SRC for the years 2014-2024. Both the number of SRCs and the percentage of SRCs within all concussions were calculated. We did not use a rate as the denominator would be the number of children and youth participating in sports which is not available particularly for sports like skiing and snowboarding. We also used the Material Deprivation Quintile from the Ontario Marginalization Index. This neighborhood-level metric consists of income, quality of housing, educational attainment, and family structure characteristics [15]. Quintile 1 represents the lowest material deprivation quintile and Quintile 5 represents the highest deprivation quintile [15]. Visits for which income quintile was missing were excluded from the analysis (<1% of visits).

An interrupted time series analysis was employed to examine changes in ED visits by policy change. Previously published research indicated that there was a rising trend in ED visits for concussion in children and youth in Ontario prior to PPM 158 from 2010-2020 [15]. The current 10 years were grouped into three time periods. The first was 2014-2018 when PPM 158 had been enacted, but prior to the enactment of Rowan’s Law. This time period served as the reference category for the time series analysis. The second time period was 2020-2021 representing the beginning of COVID-19 pandemic when many sports were not available. The third time period included 2019, 2022, 2023, and 2024 and represented the time after the enactment of Rowan’s Law excluding the pandemic years. The primary outcome variable were ED visits with a cause of injury code related to sports. This study evaluated the change in ED visits for SRCs both in total and by material deprivation quintile over a ten-year period (2014-2024). The RECORD checklist for studies involving routinely collected health services data was followed [16].

## Results

There were 70,109 children and adolescents treated for SRCs in Ontario EDs between 2014 and 2024. The percentage of SRCs within all concussions was 48.3%.

Table 1 includes the number of ED visits for SRCs and the percentage of SRCs within all concussions in children and adolescents. The number of concussions in 2024 was 807 (12%) lower than in 2014, and the percent of SRCs out of all concussions declined slightly from 52.6% in 2014 to 47.8% in 2024. Figure 2 shows the data by material deprivation quintile. Table 2 includes the Beta coefficients and p-values for the time series regression analysis for all SRCs and for SRCs by quintile. There were fewer concussions generally and by material deprivation quintile during the COVID-19 pandemic and after the enactment of Rowan’s Law, but only Quintile 1 (the highest income quintile) and Quintile 4 (second lowest quintile) were statistically significantly lower than the pre-law period.

**Table 1:**
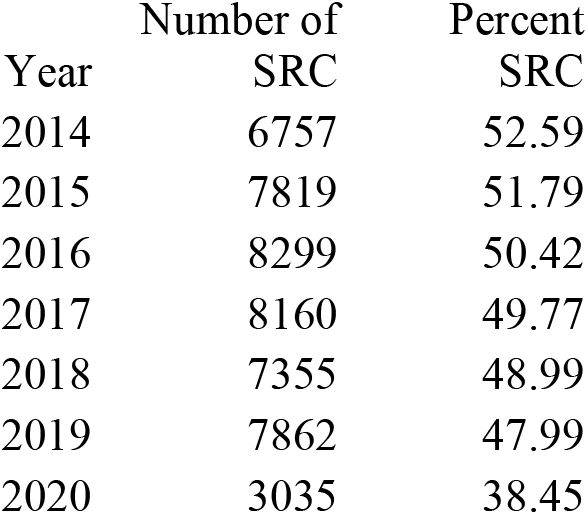
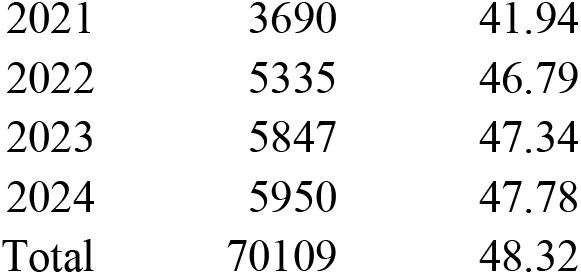
Number of SRCs and percentage of total concussions by Year, Ontario 2014-2024.

| Year | Number of SRC | Percent SRC |
| --- | --- | --- |
| 2014 | 6757 | 52.59 |
| 2015 | 7819 | 51.79 |
| 2016 | 8299 | 50.42 |
| 2017 | 8160 | 49.77 |
| 2018 | 7355 | 48.99 |
| 2019 | 7862 | 47.99 |
| 2020 | 3035 | 38.45 |
| 2021 | 3690 | 41.94 |
| 2022 | 5335 | 46.79 |
| 2023 | 5847 | 47.34 |
| 2024 | 5950 | 47.78 |
| Total | 70109 | 48.32 |

**Figure 1.**
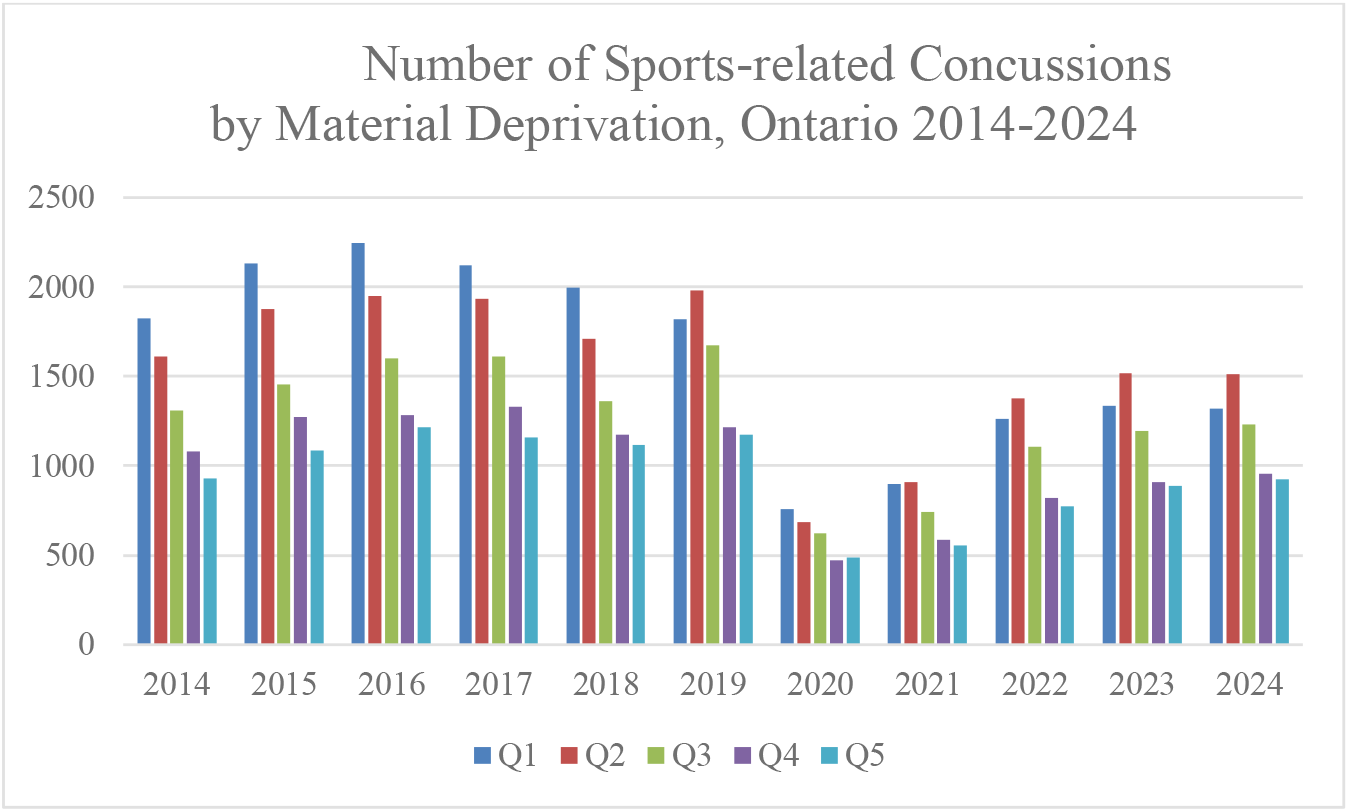
Number of Sports-related Concussions by Material Deprivation, Ontario 2014-2024

**Table 2:** Sport-related concussion Interrupted Time Series Results by Material Deprivation Quintile, Ontario 2014-2024.

| Quintile | Beta coefficient<br>Covid-19<br>(2020-2021) | p value | Beta<br>coefficient<br>post Rowan's<br>Law (2019,<br>2022-2024) | p value |
| --- | --- | --- | --- | --- |
| All | -4315.5 | <0.001 | -1429.5 | 0.033 |
| Q1 least deprived | -1332.9 | <0.001 | -628.7 | 0.001 |
| Q2 | -1019.1 | <0.001 | -218.4 | 0.144 |
| Q3 | -785.4 | <0.001 | -165.9 | 0.219 |
| Q4 | -697.9 | <0.001 | -252.4 | 0.020 |
| Q5 most deprived | -580.2 | <0.001 | -164.2 | 0.095 |

## Discussion

Sport-related concussions represented almost half of ED visits for concussions. This finding is likely due to speed and contact in sports such as ice hockey, football, and flag football [15,16]. It is worth noting that the percentage of SRCs decreased by about 5%, which means that other concussion mechanisms such as playground falls or motor-vehicle collisions remained relatively stable and unimpacted by Rowan’s Law. Further, the enactment of Rowan’s Law appears to be associated with a reduction in ED visits for SRC in Ontario generally. These reductions were statistically significant in both the least deprived income quintile (Q1) and the second most deprived income quintile (Q4). The results by material deprivation quintile suggest that children in more deprived areas are less likely to access an ED for a SRC than those in less deprived quintiles.

Since the enactment of PPM 158, all school boards are required to have a concussion policy, which includes sending information to parents about concussion tracking and treatment [4]. Coaches and sport staff have also been mandated to improve their SRC education with the help of injury prevention units such as Parachute Canada [5]. Despite these mandates, parents, coaches, and teachers in more deprived neighborhoods may have less access to such education and may experience other barriers related to language and culture [17]. Further, as the price of equipment and registration for organized sports are costly, children living in neighborhoods with less material deprivation may have a greater opportunity to engage in such sports [18]. Additionally, those from more deprived neighborhoods may be less likely to have coaches who are familiar with Rowan’s Law and PPM 158 [17]. Notably, there was a significant decrease in ED visits due to SRC in the second most deprived quintile (Q4), which suggests that the effect of Rowan’s Law went beyond the highest quintile, warranting further investigation.

### Strengths and limitations

The large population-based sample is a strength of this study as the data comes from all emergency departments across Ontario. Using ED data minimizes bias that can arise from parent or child self-report.

The primary limitation of this study is that not all sports-related concussions were captured. Those who sought care from a Sport Medicine Clinic, family physician, other health care provider, and who did not seek medical attention for a concussion were not included in this study. It is possible that referral patterns and location of follow-up services have changed because of Rowan’s Law. Subsequently, SRCs may be treated in family physician offices or sports medicine settings, neither of which were represented in the data. Finally, these policies were not enacted at one distinct point in time, and individual elements of each policy were likely spread out over the years.

## Conclusions

In conclusion, PPM 158 was associated with a rise in ED visits for concussion and the subsequent Rowan’s Law was associated with an initial decrease in ED visits for concussion that was sustained post-COVID-19 pandemic. This highlights the importance of including policy interventions to prevent and detect SRC in children and youth.

## Acknowledgements

This study was supported by ICES, which is funded in part by an annual grant from the Ontario Ministry of Health (MOH) and the Ministry of Long-Term Care (MLTC). This document used data adapted from the Statistics Canada Postal Code ^OM^ Conversion File, which is based on data licensed from Canada Post Corporation, and/or data adapted from the Ontario Ministry of Health Postal Code Conversion Fil, which contains data copied under license from © Canada Post Corporation and statistics Canada. Parts of this material are based on data and/or information compiled and provided by CIHI. The analyses, opinions, and statements expressed herein are solely those of the authors and do not reflect thos of the funding or data sources; no endorsement is intended or should be inferred. This work was supported initially by a CIHR Chair in Child and Youth Health Services and Policy Research (RN162202-278772). We thank the Toronto Community Health Profiles Partnership for providing access to the Ontario Marginalization Index. None of the authors have competing interests to declare.

## Contributorship

AKM and LR designed the study and obtained the data. MS was involved in drafting and editing the manuscript. CAE was involved in several discussions and presentations of the data and contributed to the final manuscript. SC and PF provided content related to Rowan’s Law and PPM 158. All authors approved the final version of the manuscript. AKM is the guarantor for this manuscript.

## Data availability statement

The dataset from this study is held securely in coded form at ICES. While legal data sharing agreements between ICES and the data providers prohibit ICES from making the dataset publicly available, access may be granted to those who meet pre-specified criteria for confidential access, available at www.ices.on.ca. The full dataset creation plan and underlying analytic code are available from the authors upon request, understanding that the computer programs may rely upon coding templates or macros that are unique to ICES and are therefor either inaccessible or may require modification.

